# Common Data Elements, Scalable Data Management Infrastructure and Analytics Workflows for Large-scale Neuroimaging Studies

**DOI:** 10.1101/2021.03.16.21253726

**Authors:** Rayus Kuplicki, James Touthang, Obada Al Zoubi, Ahmad Mayeli, Masaya Misaki, NeuroMAP-Investigators, Robin L Aupperle, T. Kent Teague, Brett A. McKinney, Martin Paulus, Jerzy Bodurka

## Abstract

Neuroscience studies require considerable bioinformatic support and expertise. Numerous high-dimensional and multimodal datasets must be preprocessed and integrated to create robust and reproducible analysis pipelines. We describe a common data elements and scalable data management infrastructure that allows multiple analytics workflows to facilitate preprocessing, analysis and sharing of large-scale multi-level data. The process uses the Brain Imaging Data Structure (BIDS) format and supports MRI, fMRI, EEG, clinical and laboratory data. The infrastructure provides support for other datasets such as Fitbit and flexibility for developers to customize the integration of new types of data. Exemplar results from 200+ participants and 11 different pipelines demonstrate the utility of the infrastructure.

## 2. Introduction

Neuroimaging studies such as ABCD, ADNI, Human Connectome, and Tulsa 1000 studies are significant contributors to the rapid growth of big data (Leow et al., 2009; Van Essen et al., 2013; Jernigan et al., 2018; Victor et al., 2018). In addition to the usual high-dimensional data that accompany clinical studies (e.g., genetic, cellular and clinical assessments), neuroscience studies include multimodal data for the brain (e.g., MRI, Perfusion MRI [pMRI], diffusion MRI [dMRI], functional MRI [fMRI] and Electroencephalography [EEG]). The use of various data acquisition modalities and differences in studies’ experimental designs make it challenging to provide a common data architecture that would offer easy access, scalability, management and sharing, including the ability to build analytic workflows and to run large scale analyses with increasingly large numbers of subjects. Here, we propose possible solutions to these challenges and described our specific working implementation.

As a part of the Neuroscience-Based Mental Health Assessment and Prediction (NeuroMAP) Center of Biomedical Research Excellence (CoBRE) award from National Institute of General Medical Sciences (NIGMS/NIH), the NeuroMAP Research Core provides research infrastructure to conduct advanced neuroscience research and also is responsible for providing active data management and analysis support, which includes standardization of all acquired data. Data collected for NeuroMAP consist of a core baseline assessment as well as subsequent individual projects sharing various common data elements. The research core protocol contains neuroimaging (e.g., MRI/pMRI/dMRI/fMRI/EEG), behavioral, self-report, biomarker, and actigraphy data acquired from large cohorts of participants who are then enrolled in the various other projects. Ongoing human recruitment into the core protocol is roughly 100 participants per year in phase I (five years, with a possible extension to 10 years), so that this cohort is anticipated to reach 400+ participants. Currently at year 3, 310 participants have been enrolled. A large and growing cohort size combined with several acquisition modalities amounts to a large and increasing set of heterogenous and complex data.

Large-scale data collection pipelines are complex to establish while maintaining standardized experimental protocols on both the data-acquisition hardware level and on the clinical data management level. Follow-up analyses also require further standardization, which is often implemented in ad hoc software systems at different institutions and may even vary between labs within an institution. Home-grown solutions can work adequately, and over the past decade we have collected neuroimaging data from thousands of individuals using our own internal solutions. However, in recent years, progress has been made in the scientific community toward consensus solutions to improve data management and mechanisms for data sharing (Gorgolewski et al., 2016).

There are a number of substantial costs when using custom data management solutions, not the least of which is developing the data processing standards, which can be difficult for researchers without informatics training. Idiosyncratic naming conventions and directory structures also add overhead when sharing datasets and analysis code that was developed for specific file structures. For example, a researcher unfamiliar with a particular dataset would need to learn about its conventions along with the details of the study. Sometimes, the first thing researchers do when working with a new dataset is reformat it to match a form they are familiar with, which is extra effort that could be avoided if standard formats were used. Similarly, reusing analysis code (e.g. scripts and software) often requires either extensive reworking to be compatible with a new dataset, or reformatting the target data to be compatible with the existing code.

One possible solution is the development of a complex data management system used to store, access, and even analyze neuroimaging and associated data. There have been several projects to produce such extensive systems over the past 15 years (Marcus et al., 2007; Keator et al., 2008; Van Horn and Toga, 2009; Ozyurt et al., 2010; Das et al., 2011; Scott et al., 2011; Book et al., 2013); however, they can come with significant overhead in installation, maintenance, and user training. In fact, our institute spent considerable time and resources attempting to implement one of these systems, a project which we ultimately abandoned due to excessive cost and technical difficulties.

One of the main challenges is the need for a commonly accepted data structure format that would provide a consistent and standardized way to organize multi-level neuroimaging data. The Brain Imaging Data Structure (BIDS) (Gorgolewski et al., 2016) was introduced in 2016 and promises to alleviate some of the difficulties in organizing, documenting and sharing data and code while maintaining a simple, intuitive structure that is easy to understand and work with. With metadata stored directly on disk, either in the form of file names and locations or associated JSON sidecars, BIDS avoids requiring overly complex management software or databases. The BIDS format is remarkably similar to our internally developed neuroimaging data organization solution and we decided to transition to BIDS for the NeuroMAP studies, common data elements and all new projects going forward. Wide acceptance of BIDS provides standardization across other datasets and facilitates sharing with the scientific community.

## 3. Methods

### 3.1. Data Management Infrastructure Design

The Common Data Elements and Scalable Data Managing Infrastructure can integrate neuroimage data with various other data types (Fig. 1). The CDE data are in general composed from multimodal MRI, fMRI, EEG, physiological recordings, behavioral measures, self-reports measures, actigraphy from wearable devices, and biospecimen samples (e.g., blood and microbiome). Full details of the NeuroMAP core multilevel data collection are included in Supplementary Materials. All original data sources (left side of Fig. 1) are processed and stored in order to produce a BIDS-compliant dataset (right side of Fig. 1). The middle part of the figure shows intermediate steps and storage, while the right shows the final BIDS dataset. BIDS conversion of each element is described in detail in section 3.2. Colors are used to show which raw data and samples correspond to particular elements of the BIDS dataset in its final form.

**Figure 1.**
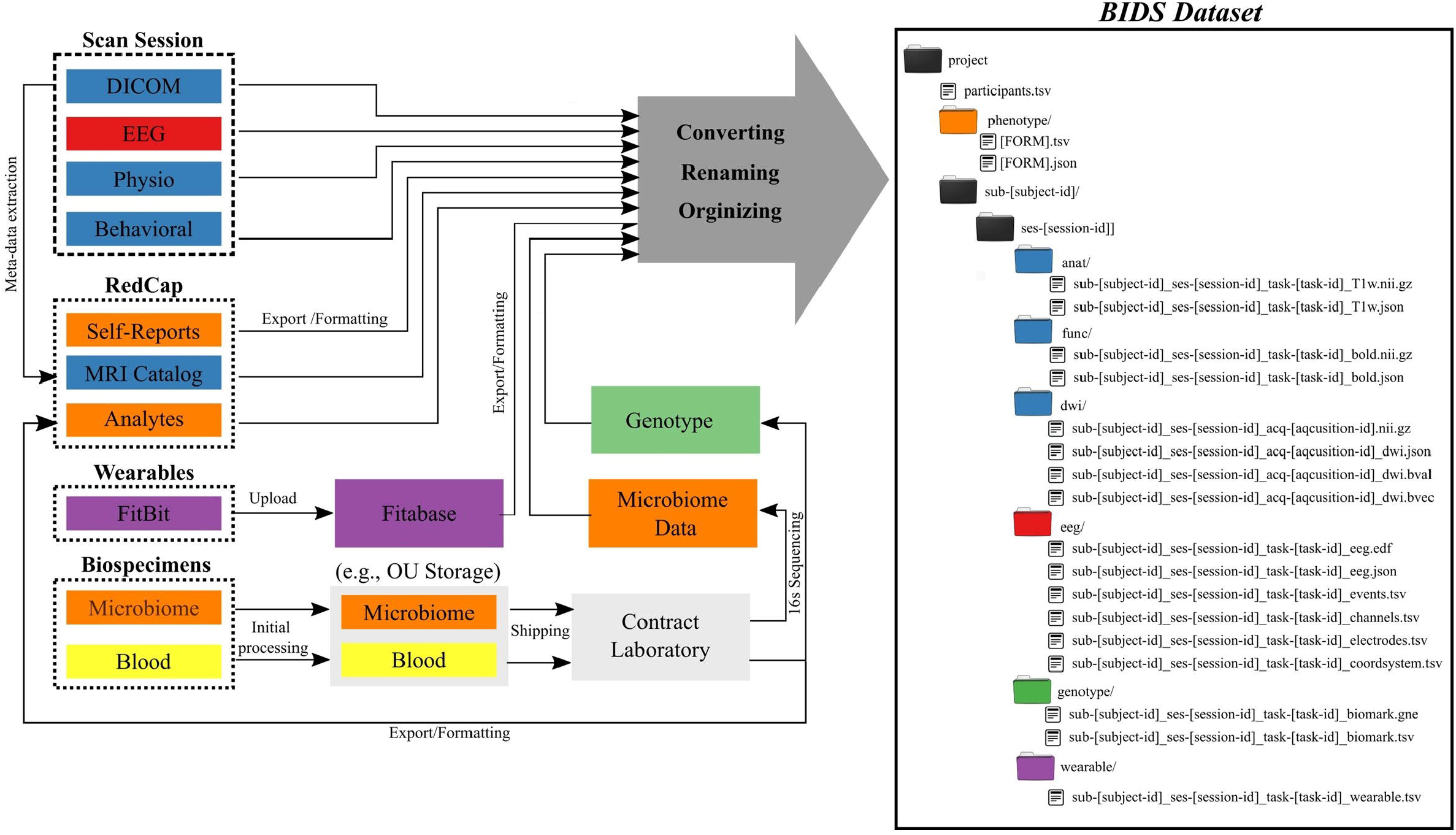
Common Data Elements and Scalable Data Management Infrastructure. Data generated and represented with different colors (left) are converted into the BIDS file structure (right), where colors of directories correspond to data types on left.

### 3.2. BIDS Conversion

#### 3.2.1. Self-report/REDCap

Self-report and clinical measures (described in full in section S-1.1.1) stored in REDCap are exported into a BIDS-compliant format using the PyCap library built on top of the REDCap API. The inputs/outputs of this process appear in orange in Figure 1. In brief, an API key links a user and access rights to a single project. Data returned from REDCap include a table of subject data for the project as well as metadata about the project and data collection instruments. The data are converted to tsv format and stored in the phenotype folder following BIDS specification. Similarly, the metadata describing the data collection instruments are stored in JSON formatted data dictionaries. The result is a json/tsv pair for each REDCap form. This script can be setup for other redcap projects and is available on GitHub (https://github.com/laureate-institute-for-brain-research/redcap-to-bids).

#### 3.2.2. Neuroimaging and associated physiological data

Neuroimaging data are produced in two formats. Source DICOM images are reconstructed and generated by the scanner and permanently stored in a read-only central location. The default organization from GE DICOM file structure has each scan stored three-folders deep (e.g., pXXX/eYYY/sZZZ, where p, e, and s refer to patient, exam, and series). For each completed scan and patient exam, these DICOM images are automatically extracted, transferred to scanner-dedicated local storage and reorganized by custom developed real-time MRI scanner data management software. To reduce the storage burden associated with hundreds of thousands of individual files, DICOM folders are packaged in .tar.gz format at the exam directory level. This reduces the number of individual files stored by a factor of 10^5^, and also saves significant storage space when individual files are smaller than the storage block size. Each DICOM image contains standard metadata indicating the subject ID, date, study, scan, and various imaging parameters: everything necessary to associate a scan with its final BIDS-compliant name and location. However, parsing through the DICOM folders and extracting metadata is an expensive operation, even before considering the compressed format. We solved this problem by creating a REDCap project called the MRI Catalogue, which contains all relevant DICOM metadata. New DICOM images from MRI scans are processed and metadata describing them are imported into REDCap nightly. Our real-time MRI software also produces a unique exam folder (on scanner-attached and dedicated real-time processing Linux workstations), which contains AFNI formatted imaging data that are uploaded and created in real time from a given session, along with any associated concurrent physiological recordings (pulse oximeter, respiratory belt, pre-processed EEG), electronic documentation for each scan with imaging parameters, DICOM file count and location on the local storage after extraction from the MRI scanner host computer and image database.

Raw EEG data (without any preprocessing) acquired concurrent with fMRI are initially stored locally on a dedicated EEG recording computer and then synchronized and transferred to network storage nightly. Similarly, behavioral responses collected during scanning tasks are initially stored on a stimulus laptop and then moved to network storage immediately upon session completion. The decision to store data locally first, then move it to network storage was based on reliability and latency considerations, so that networking issues do not affect data collection.

Neuroimaging and associated physiological data are organized and converted to BIDS format by a nightly batch process. This process handles the neuroimaging and behavioral data separately. In the first step, an export of all current MRI Catalogue data necessary for organization is extracted from REDCap. The organization process parses through these data looking for project and scan IDs matching lists for a particular project. Newly acquired matching scans are converted to nii.gz format and sent to the appropriate BIDS folder with an associated JSON sidecar. Importantly, the DICOM metadata also contains a pointer to the appropriate exam folder and series number, which is used to extract the associated physiological data. Technical issues often make data collection imperfect, e.g. scans may be aborted/restarted due to participant discomfort or imaging artifacts. Therefore, quality checks take place to help maintain data fidelity. The two most relevant checks include subject and duration matching. REDCap contains a list of subjects who have been consented for each study, so any subject ID in the MRI Catalogue that does not match a consented subject for the study in question is not included. This happens, for example, with technical scans, which should not appear in the final dataset. The case where scans are repeated, producing multiple scans of the same type is handled by matching on expected duration. Any scan that does not have the expected duration is discarded, since shortened duration indicates an incomplete scan.

The second part of the organization process handles new behavioral data found on network storage. These data are stored in a folder unique to the study and completion date/time of the session. Each behavioral folder should contain data from one subject at one visit, and any folders that contain multiple subjects or visits generate an error and are skipped until they are manually corrected. Reformatting raw behavioral data involves converting from csv to tsv, creation of a new header, and then placement in the final BIDS data structure. Raw EEG data are named according to subject ID, and quality control involves matching on subject ID, date/time, and duration, similar to what is done for imaging data.

#### 3.2.3. Behavioral data

Data management for behavioral sessions completed outside the scanner mirrors that for the behavioral data from scanning sessions, where raw files are initially stored locally, moved to network storage at the end of the session, and then parsed/organized nightly. The behavioral session also includes physiological data acquired using Acknowledge software (BIOPAC Systems, Inc.). These data are initially stored as a single continuous file in .acq format covering the entire session. Bioread (https://github.com/uwmadison-chm/bioread) is used to convert to plain text format, which is then sliced into and saved as individual tsv.gz files for each task and run. Synchronization is done using the parallel port, with a unique code indicating the start and end of each task. The appropriate header values are also extracted and stored in a JSON sidecar to be BIDS compliant.

#### 3.2.4. Biospecimens

A detailed description of initial processing and storage of biospecimens is in the supplement (S 1.1.5). Final processing of the collected samples may be carried out by a contract laboratory or done in-house and produces datasets of varying size. Blood samples are used to quantify a limited number of analytes (e.g. less than 50) describing inflammatory and metabolic states. These data are parsed and imported into REDCap for permanent storage, and then later exported into BIDS format in the same way as self-report scales. Blood samples are also sent for genotyping, which produces 650,000 or more values per participant. These data are not suitable for storage in REDCap, so they are stored in a separate repository where the location and genetic descriptors are identified in the BIDS data description. Microbiome samples produce similarly large datasets through 16S sequencing or other technologies, which again are identified in the data description to be BIDS compliant and do not have permanent storage within REDCap.

#### 3.2.5. Actigraphy/FitBit

FitBit data are initially stored in a third-party database (Fitabase https://www.fitabase.com/, accessed 2/18/2021), which handles most of the overhead related to FitBit account creation/management and aggregation of many participants’ data. Data exported from Fitabase may be divided into daily summaries and momentary assessments. Due to account management details, daily summary data often include time periods outside of the assessment windows for each subject. Start and end dates, entered into REDCap by the researcher deploying the FitBit, are used to trim the summary data down to the appropriate timeframe.

These daily summaries are stored in a single table under the phenotype folder and include overall activity levels, sleep duration and quality. Momentary assessment data including minute-wise heart rate estimates are stored in each subject’s wearable folder and are in many ways similar to behavioral outputs. Fitabase provides FitBit data in four different time intervals: 30 seconds, 1 minute, 1 hour, and 24 hours. 30-second interval data only includes sleep stages. Minute interval data include calories burned, activity intensity, metabolic equivalent of tasks (METs), current sleep stage, heart rate, and number of steps. One-hour interval data include calories burned, activity intensity, and number of steps. 24-hour interval data include activity summaries, calories burned, number of steps, and sleep.

### 3.3. Analytic Workflows

Along with the conversion of raw data into BIDS format, the Research Core also provides a set of analysis pipelines, training, and support.

#### 3.3.1. Environment

All data and analyses are hosted and completed on-site, providing full control of the systems’ configuration and operation. Our specific implementation of the primary data storage is accomplished using a network attached storage cluster running the open-source Ceph file system (CephFS). We would like to note that any modern storage hardware/solution and/or mixed local storage with cloud storage should provide alternative option for another site implementation. We selected CephFS as a scalable solution installed on commodity hardware, which allows administrators to add storage incrementally without rebuilding the entire cluster like some other solutions require. Performance scales with the size of the cluster, as data are not accessed through a fixed set of head nodes. LIBR currently has 2 petabytes of raw storage, which is 1PB of usable space after data duplication. Additionally, there is a full off-site backup copy stored roughly 100 miles away on an identical Ceph cluster. As a final precaution, LIBR also sends periodic tape backups to Iron Mountain using a Spectra BlackPearl appliance.

LIBR has 8 high-performance servers configured with the slurm workload manager (https://slurm.schedmd.com/). Each server has 24 physical cores, allowing up to 192 jobs to run in parallel and a total of over 24,000 GFlops/second. Jobs optimized to run on GPUs can take advantage of 4 Nvidia Tesla P100 cards, providing an additional 75,200 GFlops/second of computing power. Nodes are configured with 187 or 376 GB of RAM and overall networking throughput is 320 Gbps. This centralized processing infrastructure helps mitigate the bottleneck associated with network attached storage by providing 40 Gbps connections, which far outperform standard 1Gbps connections used in modern ethernet.

The storage and computing infrastructure just described was designed and developed incrementally to balance cost with performance, security, and overhead for training and maintenance. As we noted above, our data organization and processing workflows, however, do not depend on the physical details of our environment and could be implemented on a variety of systems or in the cloud.

#### 3.2.2. Pipeline Architecture Overview

Subject and group level analyses are conducted separately. Processing pipelines are implemented to service an individual subject and analysis, where an analysis typically deals with one task and set of processing parameters (Fig. 2). This allows for parallelization at the subject plus pipeline level, with separate jobs submitted for each subject.

**Figure 2.**
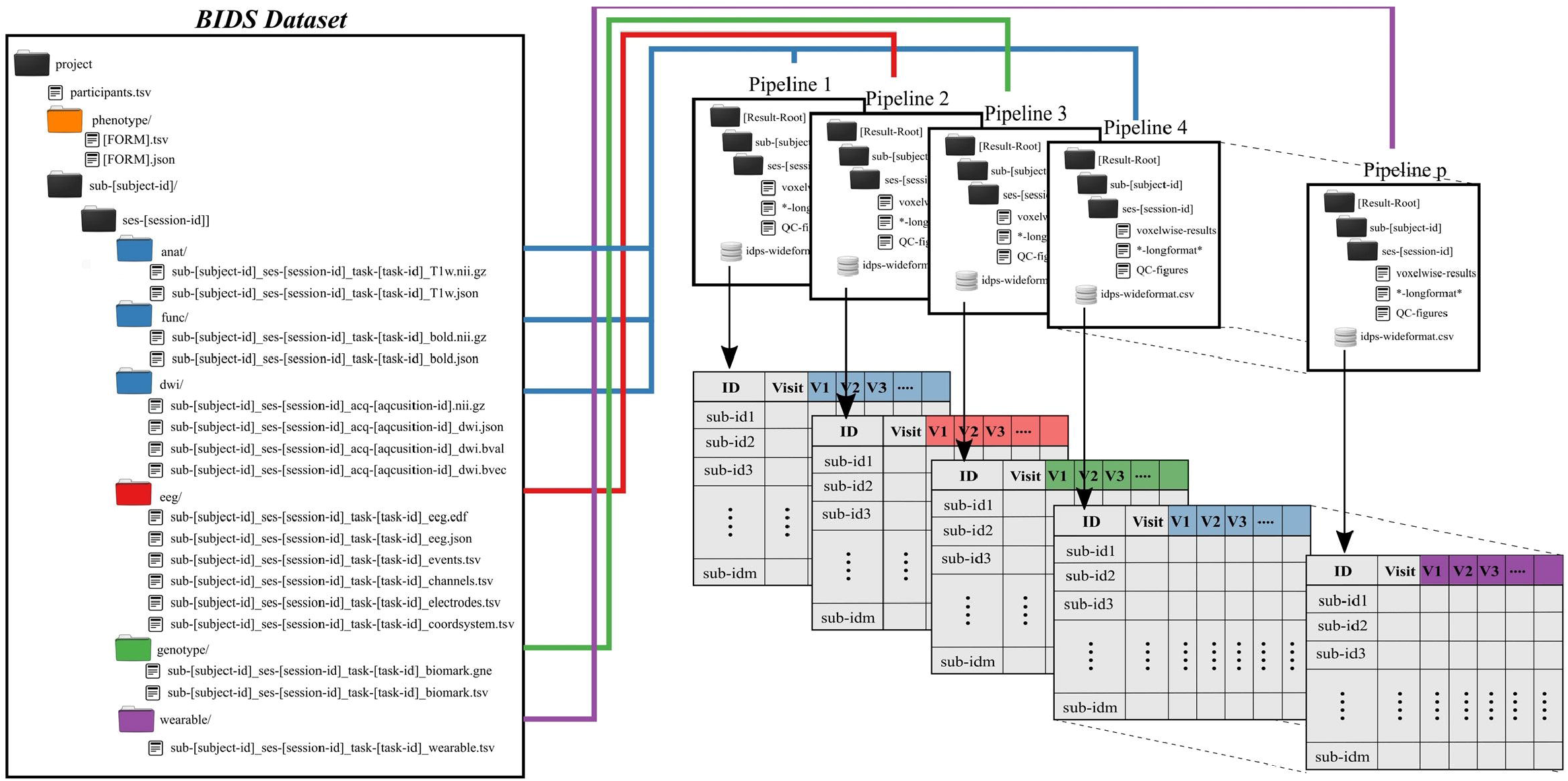
Preprocessing pipelines operate on BIDS-formatted inputs and create output in tabulated form for group level analysis. Derived data are colored to match raw data sources.

All single-subject analyses are submitted to the batch scheduler using a script named preprocess-all-BIDS.py. This wrapper reads in a configuration file with pointers to the root of the source data directory (i.e., the root of one BIDS dataset), the desired root of the output directory tree and which pipeline to run. The output directory structure mirrors the BIDS formatted input, so that individual subject/session/pipeline results are stored in [Results Root]/sub-[subject]/ses-[session]/[leaf]. preprocess-all-BIDS.py traverses the input folder structure, and for every subject/session checks to see if a job has already been submitted, based on the existence of specially named status-indicating files in the output directory. If this subject/session combination has not been run for this pipeline, the output directory is created and a job is submitted.

Results for an individual subject/session/task/pipeline include derived values to be tabulated, quality control images in png or jpg format, and larger format derived data, like voxelwise statistics. Derived values include metrics like subject head motion, subject performance including mean reaction times and accuracy, physiological measures including heart rate, and in the case of imaging tasks, extracted activations, contrasts, and volumes from atlas-based regions of interest. All derived values are stored in files ending in the .longformat suffix, where these are simple text files in attribute-value format. After processing data for all subjects, all values found in .longformat files are combined, producing a consolidated table with a single row per subject and session and one column per attribute. This consolidated format, ready for use in various statistics applications, is saved as in .csv and, RData formats, the later binary being preferable for large imaging datasets with tens of thousands of variables, which can lead to performance issues when reading in text data.

Any manual quality control processes are simplified by storing appropriate images in jpg or png format. For examples, this may include EKG traces with identified R wave peaks, or montages showing alignment and normalization of neuroimaging data. This allows the user to flip through QC images for a dataset relatively quickly without, for example, needing to open neuroimaging data in specialized software.

#### 3.3.3. Neuroimaging Pipeline Options

##### fMRI Pipelines

Neuroimaging processing pipelines necessarily include numerous decisions, such as which software to use, whether to include linear or non-linear normalization to standard space, what smoothing kernel to apply, what nuisance regressors to use at the regression step and so on. These analysis decisions can impact the final results and interpretation of a study, which was recently illustrated through divergent results obtained by 70 independent groups of researchers who all analyzed the same data (Botvinik-Nezer et al., 2020). Therefore, frameworks like ours that allow the sharing of analysis workflows are essential for reproducibility and replicability. An individual researcher may customize a particular pipeline or use one of our 3 standard options for each fMRI task, labeled P01 through P03. P01 is a traditional approach using AFNI (Cox and Hyde, 1997) and includes removal of the first 3 volumes, despiking, slice-timing correction, co-registration between functional and structural volumes, motion correction, 4mm of gaussian blur, and an affine transformation to standard space. P02 is similar to P01, except that it includes a non-linear warp to standard space and RETROICOR correction (Glover et al., 2000), which helps remove physiological noise but requires the collection of pulse oximeter and respiratory belt data.

P03 takes a completely different approach, instead using fMRIPrep (Esteban et al., 2019) to do all preprocessing up until the regression step, which still uses AFNI’s 3dDeconvolve. Preprocessing with fMRIPrep uses mainly default parameters, so that a combination of tools are used to 1) select a reference fMRI volume (mean of high contrast available in initial pre T1-saturation or pre Steady State Free Precession fMRI volume); 2) perform boundary based registration with the T1-weighted images (Greve and Fischl, 2009); 3) estimate head motion prior to any spatiotemporal filtering using mcflirt in FSL 5.0.9 (Jenkinson et al., 2002); 4) perform slice timing correction using AFNI (Cox and Hyde, 1997); 5) perform nuisance regression including regressors for Framewise Displacement and DVARS (Power et al., 2014); average CSF, white matter, and whole brain signals, as well as physiological regressors using CompCor (Behzadi et al., 2007). Regardless of the pipeline, standard derived data from task-based fMRI include regression coefficients and contrasts extracted for each ROI in several atlases and summaries of head motion for quality control.

Resting state preprocessing P04 pipeline includes the same options as task data, with the addition of a fourth option, which is similar to P02 pipeline but also includes additional motion correction prior to slice timing correction via an automatic EEG assisted slice-specific motion correction for fMRI (aEREMCOR) (Wong et al., 2016). While it would be possible to include this additional motion correction step for task-based data, it is particularly important in resting state, where the residual effects of head motion are well known, and they might differ for each acquired slice (Power et al., 2015). Standard derived data from resting-state fMRI include a correlation matrix between pairs of ROIs from multiple atlases (e.g. the Brainnetome (Fan et al., 2016)) and summaries of head motion.

##### EEG pipelines

Simultaneous EEG-fMRI offers several benefits to measure and study the human brain’s spatial and temporal dynamics in health and disease. However, EEG data collected during fMRI acquisition are contaminated with MRI gradients and ballistocardiogram artifacts, in addition to artifacts of physiological origin (eye blinks, muscle, motion), these artifacts need to be detected and suppressed before further data analysis (Mayeli et al., 2019). We have developed in house a comprehensive automated pipeline for EEG artifact reduction (APPEAR) recorded during fMRI, which we have incorporated into the BIDS preprocessing pipeline architecture (Fig. 2). APPEAR is capable of reducing all main EEG artifacts, including MRI gradients, BCG, eye blinks, muscle, and motion artifacts, and can be applied to large (i.e., hundreds of subjects) EEG-fMRI datasets. APPEAR was evaluated, tested and compared to manual pre-processing EEG data for both resting EEG-fMRI recording as well as for event-related potential or task-based EEG-fMRI experiments in an exemplar eight subject EEG-fMRI dataset.

## 4. Results

We provide examples illustrating pipelines P01 through P04 to help demonstrate the utility of the processing infrastructure.

### 4.1. Task fMRI Results

Exemplar CDE data have been processed for the Monetary Incentive Delay and Stop Signal tasks (see the supplementary material for tasks details). Figure 3a shows voxel-wise maps for the P5 - P0 contrast in the MID as produced by pipelines P01 through P03. Data from 93 participants are included here, with pipelines P01 through P03 taking approximately 1.3, 4, and 5 CPU hours per subject to complete. With the architecture detailed in 3.3.2, processing for all three sets of data could be completed in under one day when all resources are available. The alignment QC images produced by each pipeline make it possible to complete all manual QC for roughly 100 participants and one pipeline in less than one hour. Figure 3b shows voxelwise maps of the Stop – NoStop contrast from the stop signal task, again produced by pipelines P01 through P03. These maps include data from 49 participants.

**Figure 3.**
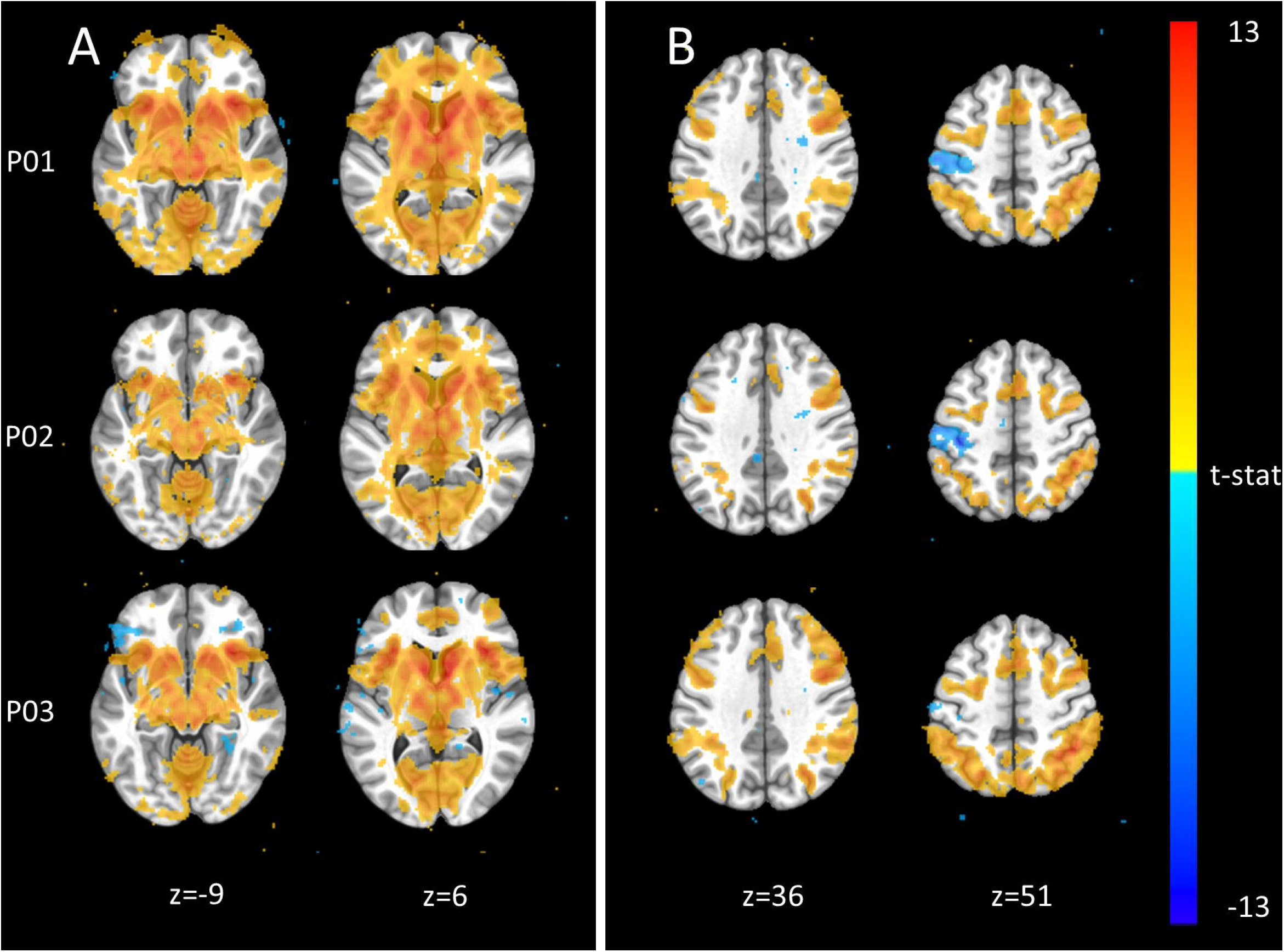
Exemplar voxel-wise task activation maps produced by three different pipelines. A) Monetary Incentive Delay P5 – P0 contrast from n=93 participants at p < 0.001. B) Stop Signal Stop – NoStop contrast from n=49 subjects at p < 0.001.

### 4.2. Resting State fMRI Results

Exemplar CDE data have also been processed for resting-state fMRI using all four pipelines, P01-P04. Figure 4a shows the average connectivity matrix extracted from the Brainnetome atlas and organized by approximate networks identified using the Yeo 7-network atlas (Yeo et al., 2011). All pipelines produce qualitatively similar results at the group level.

**Figure 4.**
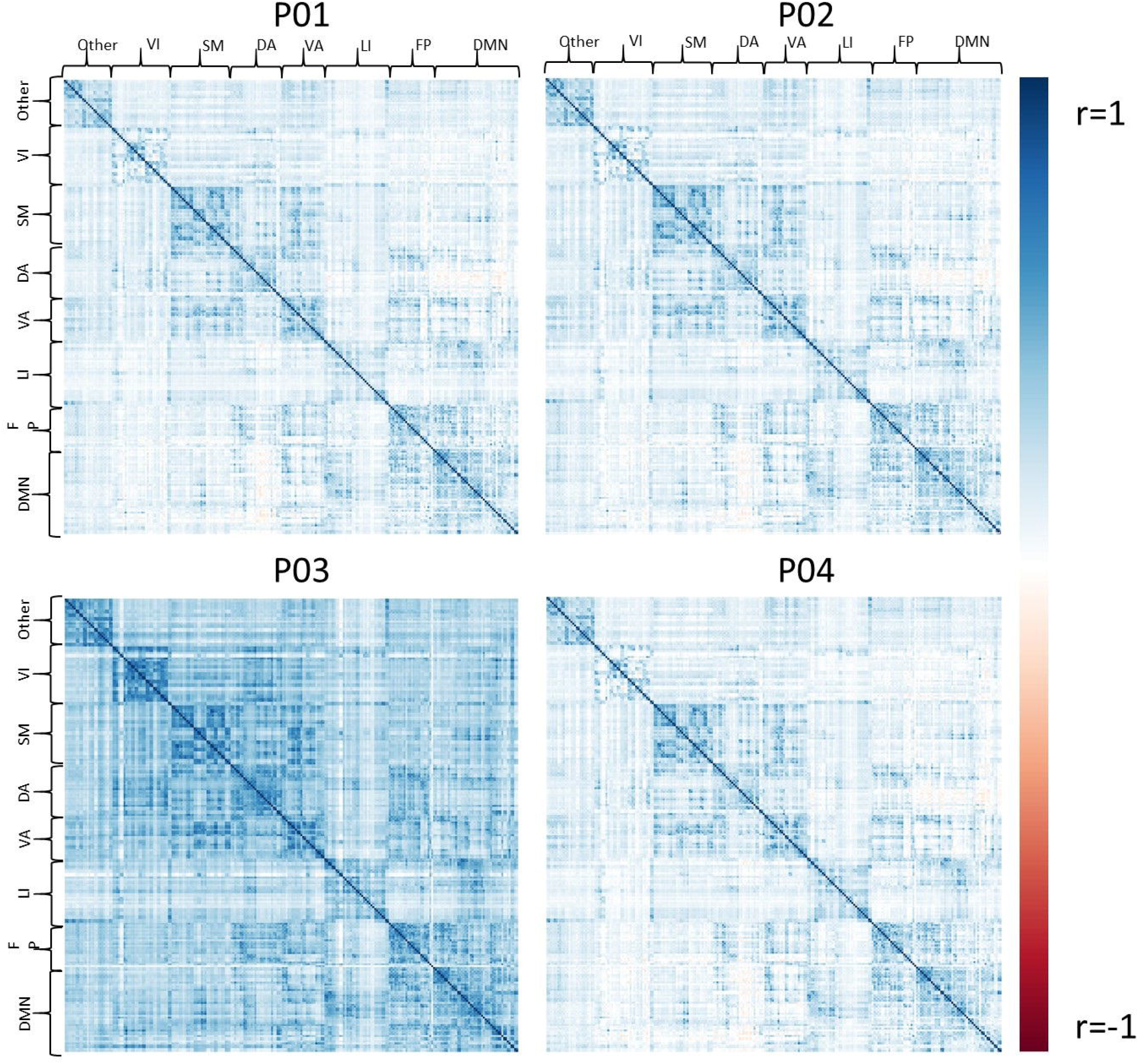
The set of group average correlation matrices from resting state with: P01 (linear registration), P02 (nonlinear registration+RETROICOR correction), P03 (fMRIPrep), P04 (P02 + aEREMCOR).

Figure 5 shows the relationship between individual features (correlation strengths) measured with different pipelines. Points on the 45 degree line indicate complete agreement between methods, while divergence from that line illustrates differences between pipelines.

**Figure 5.**
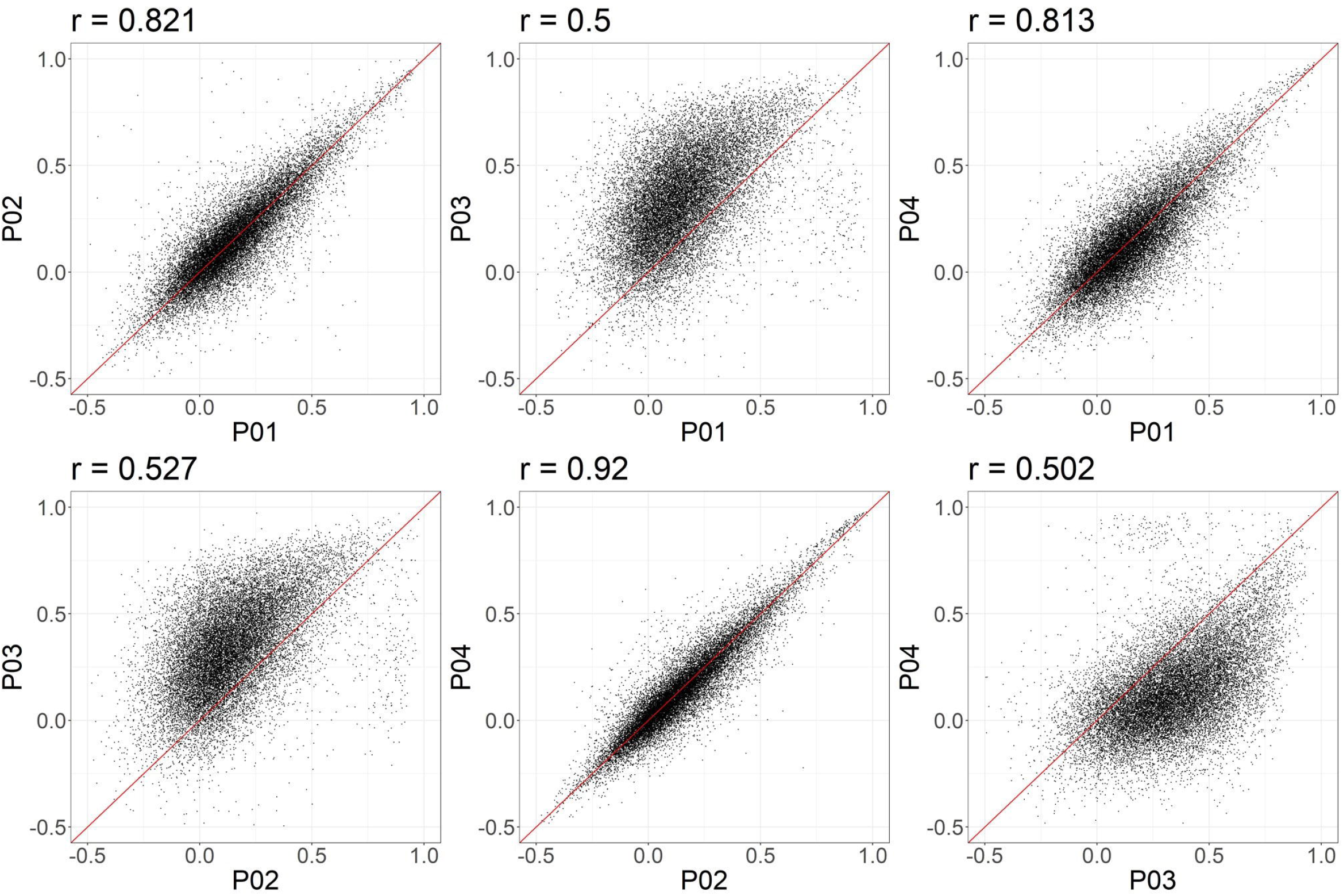
Node-to-node correlations measured for individual subjects. Each point represents the connectivity measured for one pair of ROIs and one subject, with the X and Y values representing the connectivity measured obtained with two different pipelines. 20,000 points were randomly sampled for plotting.

### 4.3. EEG Preprocessing

We have utilized the APPEAR pipeline to preprocess EEG data acquired concurrently with fMRI, and then applied comprehensive EEG feature extraction from five subsets of EEG features including amplitude, connectivity, fractal dimension (FD), range and spectral power features. Furthermore, each subset of features was applied to Alpha [8-13] Hz, Beta[15-30] Hz, Theta[4-7] Hz, Delta[0.5-4] Hz, Gamma [30-40] Hz and whole range of EEG frequency [0.5-40] Hz. An exemplar EEG feature correlation matrix is shown in Figure 6. The exemplar use of the extracted EEG features and automated EEG preprocessing can found elsewhere [https://github.com/obada-alzoubi/Comprehensive_EEG_Features_Extraction].

**Figure 6.**
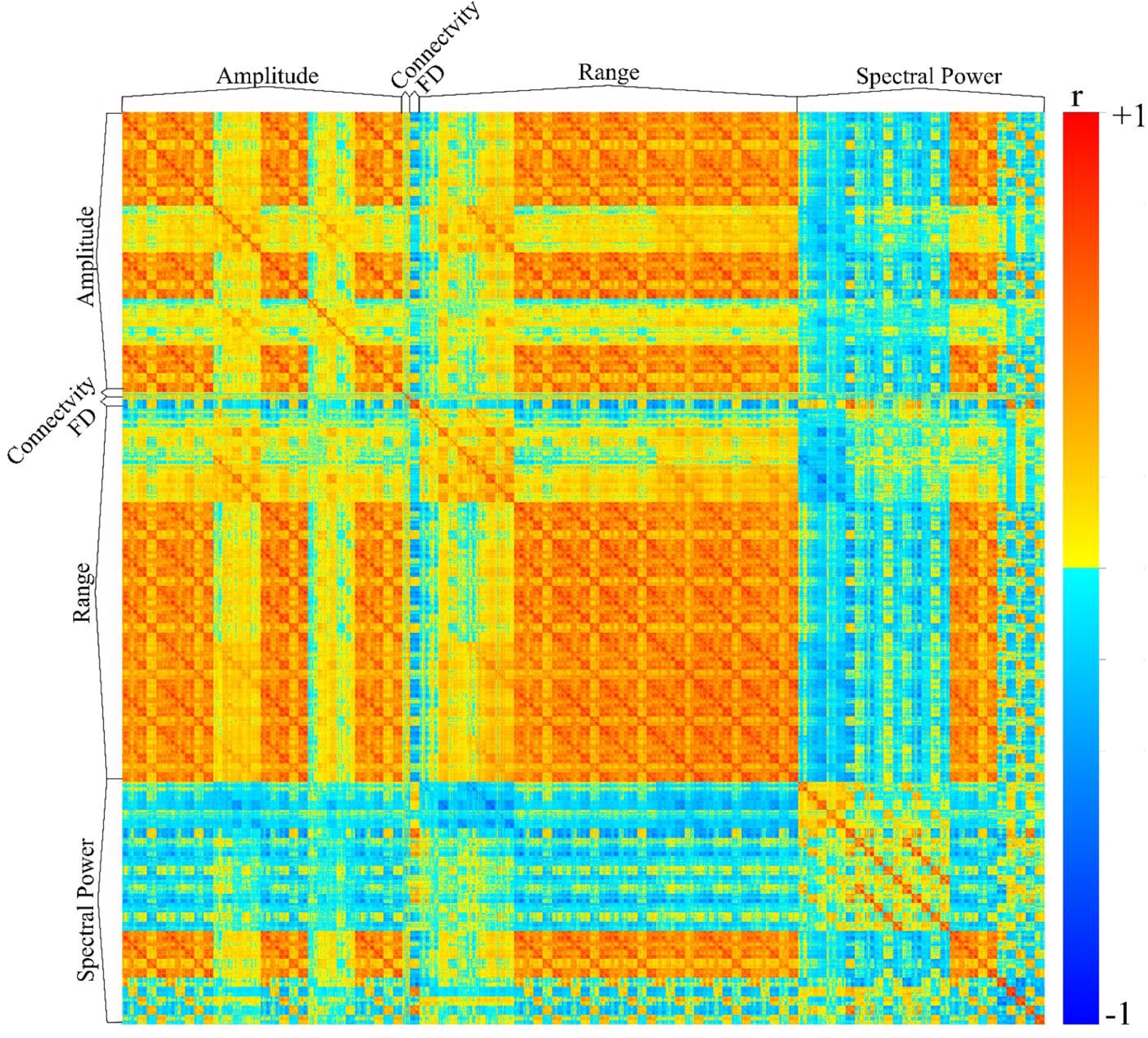
The correlation matrix of 3032 EEG features extracted using comprehensive EEG features extraction for resting-state condition. Five different subsets of features were extracted including Amplitude (31 Channels × 5 bands× 6 types = 930 features), connectivity (24 features), FD (31 Channels × 1 Feature=31), range (31 Channels × 5 bands× 8 types = 1240 features) and spectral power features (31 Channels × 5 bands× 5 types + 31 Channels ×1 Feature = 806 features). For more details about each subset of features, please see (Al Zoubi et al., 2018).

## 5.4. Discussion

The proliferation of high-throughput data-generating technologies in biomedical research has led to data analytics challenges for creating easily reusable and reproducible pipelines. These challenges are especially salient for neuroscience studies, which not only involve the usual high-dimensional data but also include multiple neuroimage-specific data types and complex psychological trait data. The current study describes a scalable environment and set of software pipelines to preprocess neuroimaging (MRI, fMRI, and EEG) and behavioral data while integrating them with other subject-level high-dimensional data to perform sharable, reproducible analyses.

The services and computational environment developed by the Research Core provide a set of tangible benefits to ongoing research. Massive amounts of complex neuroimaging data are put into a standard (BIDS) format with minimal human interaction in an ongoing basis. The architecture for converting data to BIDS format is flexible and scalable, so that new studies often have compliant data from day 1.

Once the data for a study are in BIDS format, running any of our standard preprocessing pipelines becomes a quick process. With relatively little human intervention, preprocessing jobs can be created for hundreds or thousands of participants, and the processing and network storage infrastructure can produce results in days rather than weeks. Having multiple pipelines available for the same tasks gives researchers the ability to verify that their results are robust to the details of the preprocessing pipeline, as others have shown the wide variation in analysis results to be a serious concern (Botvinik-Nezer et al., 2020).

In this work, we provide exemplar results 11 different pipelines (3 pipelines on each of 2 fMRI tasks, 4 pipelines on resting-state fMRI, and one pipeline on resting EEG) to demonstrate the utility of our infrastructure. Additionally, ROI-level results from our standard pipelines have been used in studies of cannabis (Spechler et al., 2020) and stimulant/opioid use (Stewart et al., 2020), while voxelwise results have appeared in studies of neighborhood effects (Feng et al., 2019) and inflammation (Burrows et al., 2021), and clinical data have been used to predict head motion during scanning (Ekhtiari et al., 2019). We have also used EEG derived features have to differentiate participants with mood and anxiety disorders from healthy controls (Al Zoubi et al., 2019) and to predict participant age (Al Zoubi et al., 2018).

Our workflow incorporates many diverse processing and analysis tools such as afni, freesurfer, fmriprep and uses the BIDS format. However, it has been noted that the large number of analysis degrees of freedom in neuroscience increases the risk of false discoveries due (Wicherts et al., 2016). Each analysis step can result in an expanding decision tree of potential analyses. Determining the best workflow software or pipeline option for a given experiment is an ongoing question, but the current software provides standard selections for the many analysis options. As the field evolves and standards consolidate, the default processing and analysis parameters will converge to standards with lower variation and increased replicability.

In addition to neuroimaging data, our current pipelines include other common data types and can be easily extended to other high-throughput data, such as genetic and gene expression. Many neuroscience studies also include large non-neuroimage datasets, such as GWAS, which has its own relatively complex file format known as Plink. BIDS is opensource and under active development, and integration with these other datasets will be straightforward extensions of BIDS.

## 6. Ethics and Dissemination

Human neuroimaging data were acquired as part of NeuroMap CoBRE Award from National Institute of General Medical Sciences, National Institutes of Health P20GM121312 award. The NeuroMap CoBRE Research Core IRB protocol (WIRB protocol number 20182352) was approved by the Western Institutional Review Board, Puyallup, WA. In addition all individual NeuroMap Investigators studies’ protocols were approved by the Western Institutional Review Board, Puyallup, WA. All human research was conducted according to the principles expressed in Declaration of Helsinki. All subjects gave written informed consent to participate in the study and received financial compensation.

## Data Availability

The exemplar dataset used in this study to evaluate fMRI preprocessing pipelines will be prepared and made available upon reasonable request to the corresponding author.

## Acknowledgments

We thank Dr. Jennifer Stewart for valuable training contributions for NeuroMap-Investigators. The NeuroMAP-Investigators include the following contributors: Yoon Hee-Cha MD., Justin Feinsten Ph.D., Sahib Khalsa MD., Jonathan Savitz Ph.D., Kyle Simmons Ph.D., Namik Kiric Ph.D., Maria Ironside Ph.D., Evan White Ph.D.

