## Supplement for "Common Data Elements, Scalable Data Management Infrastructure and Analytics Workflows for Large-scale Neuroimaging Studies"

Rayus Kuplicki^1#^, James Touthang^1^, Obada Al Zoubi^1^, Ahmad Mayeli^1^, Masaya Misaki^1^, NeuroMAP-Investigators^1,2^, Robin L. Aupperle^1,2^, T. Kent Teague^3,4,5^, Brett A. McKinney^6,7^, Martin P. Paulus^1^, and Jerzy Bodurka^1,8#^

^1^Laureate Institute for Brain Research, Tulsa, OK, USA

^2^Department of Community Medicine, Oxley Health Sciences, University of Tulsa, Tulsa, OK, USA

^3^Department of Surgery, University of Oklahoma School of Community Medicine, Tulsa, OK, USA

^4^Department of Psychiatry, University of Oklahoma School of Community Medicine, Tulsa, OK, USA

^5^Department of Biochemistry and Microbiology, Oklahoma State University Center for Health Sciences, Tulsa, OK, USA

^6^Department of Mathematics, University of Tulsa, Tulsa, OK, USA,

^7^Tandy School of Computer Science, University of Tulsa, OK, USA

^8^Stephenson School of Biomedical Engineering, University of Oklahoma, Norman, OK, USA

^#^Corresponding authors:

Dr. Rayus Kuplicki;

Dr. Jerzy Bodurka;

**NeuroMap-Investigators**:

The NeuroMap-Investigators include the following contributors: Yoon Hee-Cha MD., Justin Feinsten Ph.D., Sahib Khalsa MD., Jonathan Savitz Ph.D., Kyle Simmons Ph.D., Namik Kiric Ph.D., Maria Ironside Ph.D., Evan White Ph.D.

**Multi-level data collection**

The NeuroMAP Core includes common data elements collected from five general modalities. Each modality (self-report, neuroimaging, behavior, actigraphy, biospecimen) probes one or more different RDoC domains (Sanislow et al., 2010). Here we detail the measures collected with each modality along with the domains being assessed.

### 1.1.1. Self-report/REDCap

All self-reported and clinical interview data are collected directly into an internally-hosted instance of the Research Electronic Data Capture (REDCap) (Harris et al., 2009) database. REDCap is a secure (e.g. compliant with 21 CFR Part 11, FISMA, HIPAA, and GDPR), web-based data collection system that is currently used by over 3,200 institutions in 128 countries (Harris et al., 2019). We adopted REDCap in 2014 to replace a combination of paper-charts and a home-grown databasing solution. Briefly, the selected assessments draw heavily from three different sources. Wherever suitable, Patient-Reported Outcomes Measurement Information System (PROMIS) measures are used because they provide brief, computer adaptive tests and minimize participant burden (Ader, 2007). We also rely heavily on measures from the PhenX Toolkit, which contains consensus measures for **Phen**otypes and e**X**posures from 25 different domains (Hamilton et al., 2011). Any constructs that are not sufficiently captured by either PROMIS or PhenX measures are collected using standard questionnaires.

Neuropsychological testing is done using modules from the NIH Toolbox (Gershon et al., 2013), which provides a set of standard assessments administered via iPad or other mobile devices. All data collected through the NIH Toolbox are transferred to a LIBR hosted server which handles reformatting and import into REDCap. While there may be some benefit to traditional interview-style neuropsychological testing, the ability to administer and score tests with minimal staff effort, especially from bypassing any manual scoring, makes high-volume data collection more feasible.

### 1.1.2. MRI and simultaneous EEG-fMRI data acquisition

#### 1.1.2.1. Equipment and Overview

Two research dedicated 3 Tesla MRI scanners (General Electric Discovery MR750) are used for all neuroimaging research. Both scanners allowed for state-of the art MRI imaging acquisitions including massive parallel and accelerated imaging including Sensitivity Encoding, and multi-slice imaging. Each scanner is equipped with specialized MRI signal detectors (RF coils) allowing for scanning human brain, and spine at high spatial and temporal resolution and excellent image quality. Each scanner is also equipped with dedicated MR-compatible 128-channel EEG system (Brain Products GmBH), and custom build real-time fMRI system which provided advance solution for extraction, managing, conversion and storage of all neuroimaging data, and allowing for real-time fMRI, simultaneous EEG-fMRI with neurofeedback, as well simultaneous EEG-fMRI hyperscanning.

The NeuroMAP Core includes 90-minute functional MRI (fMRI) session conducted with simultaneous EEG recordings and 30-minute structural MRI scans. The two scanning sessions have slightly different hardware configurations, however, in many cases can both be performed in a single visit. When performing a full 2-hour session, participants first complete the functional session, then they are brought out of the scanner briefly while the necessary changes are made and put back in for the functional session. This allows us the flexibility either to complete both sessions in a single visit or to split it into two sessions, depending on scheduling constraints and participant preference.

#### 1.1.2.2. Functional Imaging

fMRI data are acquired using a standard 8-channel GE head coil and with SENSE accelerated single shot gradient echo EPI sequence.. All fMRI scans used the same EPI parameters (TR/TE = 2000/27 ms, FOV/slice = 240/2.9 mm, 39 axial slices, SENSE acceleration R=2, 96 × 96 matrix reconstructed into 128 x 128 image resulting into 1.875x1.875x2.9mm^3^ voxels volume) with varying numbers of TRs for different runs. Concurrent 32-channel EEG data are collected simultaneously with all fMRI experiments. Both raw and pre-processed (i.e. removal of MR and cardioballistic artifact using Brain Products’ proprietary software) data are saved. All functional imaging also includes concurrent recordings of cardiac waveform via a finger pulse oximeter and respiratory waveform using an abdominal belt, which can both be used for fMRI physiological noise correction (Glover et al., 2000). A high-resolution T1-weighted structural MP-RAGE is collected using the following parameters: slice orientation=sagittal, FOV=256mm, image matrix=256×256, slice thickness=1.0mm producing 1mm isotropic voxels, 208 slices, TR/TE=6/2.92 ms, SENSE acceleration ratio R=2 in the phase encoding direction, flip angle=8 degrees. Functional scans include resting state followed by three tasks (Visceral Interoceptive Awareness, Monetary Incentive Delay, Fear Conditioning).

##### 1.1.2.2.1. Resting State

Two six-minute (180 TRs) scans are collected with participants at rest. They are presented with a white crosshair in the center of an otherwise black screen and are instructed to “Focus on the plus-sign, clear your mind and try not to think of anything in particular.”

##### 1.1.2.2.2. Visceral Interoceptive Attention

The visceral interoceptive attention task contains two six-minute (180 TRs) runs and is adapted from (Simmons et al., 2013), which has been shown to activate sub-regions of the insula which respond differentially to interoceptive and exteroceptive signals. This event-related task contains two interoceptive (heart, lungs), and one exteroceptive (target) conditions. In the interoceptive attention conditions, participants are presented with the text “Heart” or “Lungs” as black text on a white background and are instructed to focus on sensations coming from that part of the body. On exteroceptive attention trials, participants are presented with the word “Target” in text which alternates between black and a shade of grey at a rate of one switch per second. The intensity of the color change has seven levels varying from no change (i.e. the text remains black for the duration) to near-white. During these trials participants are instructed to focus on the intensity of the color change. All trials (interoceptive and exteroceptive) are 10 seconds in duration and half of trials are followed by ratings where participants use a button box to rate the intensity of the sensations or color change on a 0 to 6 scale. There are 6 trials per condition in each run, for a total of 12 Heart, 12 Lungs, and 12 Target trials. Participants are presented with a fixation cross with a duration ranging from 2.5 to 15 seconds between trials.

##### 1.1.2.2.3. Monetary Incentive Delay

The monetary incentive delay task is adapted from (Knutson et al., 2001) and contains two 9-minute, 22-second (281 TRs) runs. Each trial in this task consists of cue presentation indicating a potential gain (circle) or loss (square), followed by a brief fixation and then display of a target (white triangle). The magnitude of potential gain or loss on each trial is indicated by the position of a horizontal line on the cue along with text and had values of 0, 1, or 5 US dollars. Regardless of condition, participants are tasked with pressing a response button while the target is displayed. Each trial ends with presentation of the outcome. There are a total of 90 trials (45 per run, 15 per condition), and task difficulty was calibrated based on subject performance during a practice run such that participants were expected to succeed on two thirds of trials, resulting in an average total reward of $30.

##### 1.1.2.2.4. Fear Conditioning

The fear conditioning task is similar to (Ball et al., 2017) and contains three phases (familiarization, conditioning, extinction) spread across four runs. Two neutral, abstract images are used as conditioned stimuli (CS) and are randomly assigned as either CS+ (to be paired with an aversive unconditioned stimulus US) or CS-, not paired with the US. The US is a 1 second aversive scream beginning 500ms after image onset for a subset of CS+ trials. CS presentations are performed in a pseudorandomized order and between US presentations participants complete a continuous performance task where they press a left or right button in response to left or right facing arrows, which helps maintain task engagement. The familiarization phase consists of a single 2-minute 38-second (79 TRs) run with 5 presentations of each CS without any US presentation. The conditioning phase is split into two 8-minute 40-second (260 TRs) runs each containing 15 presentations of each CS without the US and 5 presentations of the CS+ paired with the US. The extinction phase consists of a single 12-minute 16-second (368 TRs) run containing 25 presentations of each CS with no US presentation. Participants also make valence, arousal, and anxiety ratings in response to each CS at the end of each run.

#### 1.1.2.3. Structural Imaging

The structural imaging session provides a brief (30 minute) assessment including scans based on four different anatomical contrasts. This session is acquired using a 32-channel receive only array (Nova Medical, Wilmington MA). A T1-weighted MP-RAGE is collected with the following parameters: slice orientation=sagittal, FOV=256mm, image matrix=256×256, slice thickness=1.0mm producing 1mm isotropic voxels, 208 slices, TR/TE=6/2.92 ms, acceleration factor R=2 in the phase encoding direction, flip angle=8 degrees. Other than the head coil, these parameters exactly match the T1 weighted scan collected as part of the functional session and will allow us to measure short-term test-retest reliability of structural measures. In addition, matching T1-w scan prescription, a T2 weighted scan is collected with the following parameters: slice orientation=sagittal, FOV=256mm, image matrix=256x256, slice thickness=1mm producing 1mm isotropic voxels, 204 slices, TR/TE=3309/73 ms, acceleration factor R=2 in the phase encoding direction. The T1 and T2 weighted images are acquired in the same grid, which will allow us to create combined contrast images (CITE). A T2-weighted Fluid Attenuated Inversion Recovery (T2-FLAIR) is acquired with the following parameters: slice orientation=axial, FOV=240mm, image matrix=512x512, slice thickness=4mm with 1mm gap producing 0.469x0.469x5mm voxels, 30 slices, TR/TE=8000/126.7ms. Multi-band, multi-shell diffusion tensor imaging (DTI) is also acquired with the following parameters: 102 diffusion encoding directions, b values = 500, 1000, 2000, and 3000 s/mm2, FOV=240mm, acquisition matrix=140x140, slice thickness = 1.7mm producing 1.714x1.714x1.7mm voxels, 80 slices, TR/TE=4100/81.7ms, 12 non-diffusion weighted images. Another 53 images with the phase-encoding direction reversed.

### 1.1.3. Behavioral Session

#### 1.1.3.1. Equipment and Overview

The NeuroMAP core includes a 1.5 hour behavioral session with six different computerized tasks with concurrent physiological and facial recording. All physiological recordings are collected using a BIOPAC MP 150 system and AcqKnowledge software (Lehigh, Pennsylvania). These concurrent recordings will allow a more thorough understanding of the function of the sympathetic and parasympathetic nervous systems during decision making, stress, and emotionally salient stimuli.

Galvanic skin conductance (GSR) is recorded using electrodes placed on the palmar surface of the non-dominant hand and prepared with isotonic (0.5% saline) paste. GSR measures variations in sweat gland activity and is considered to be a broad, non-valence-specific measure of emotional arousal (Greenwald et al., 1989). GSR has been shown to vary with different levels of trait anxiety and social desirability, and may predict vulnerability to anxiety disturbances (Najstrom and Jansson, 2006).

Cardiac measures are collected using 3-lead electrocardiogram (ECG), with electrodes placed below the right clavicle, below the left rib, and using the GSR electrodes as a shared ground. ECG is used to measure heart rate, which tends to increase under states of high emotional arousal. Respiratory Sinus Arrythmia is used to describe dynamic changes in heart rate related to breathing and vagal tone, which may be abnormal among those with various anxiety disorders (Watkins et al., 1998; Licht et al., 2009; Miu et al., 2009).

Respiration rate is measured using BIOPAC’s Respiration Transducer, which uses a belt wrapped around the participant’s thorax that responds to changes in chest circumference as the participant breathes. While primarily regulated by metabolic demands, respiration is also impacted by emotional state, particularly by activity of the piriform-amygdala complex (Homma and Masaoka, 2008).

Facial video is also recorded during the behavioral session as another non-invasive measure of emotional reactivity. Facial expression due to autonomic nervous system activity reflects emotional state (Ekman et al., 1983). Advances in computer vision and machine learning allow for facial recognition (Bartlett et al., 2002) and action unit coding (Donato et al., 1999; Wu et al., 2012). Emotional state can be detected either from these action units (Lien et al., 1998) or by passing the original image through a combination of Gabor filters and support vector machines (Susskind et al., 2007).

### 1.1.4. Behavioral Tasks

#### 1.1.4.1. Handgrip strength and endurance

While handgrip strength has been associated with all-cause mortality (Celis-Morales et al., 2018), handgrip endurance, which can be measured with high reliability (Gerodimos et al., 2017), may index motivation. Both are measured using an analogue dynamometer from Baseline Evaluation Instruments that includes an electrical output that recorded through the BIOPAC system. The handgrip strength protocol is adapted from the NiH Toolbox (Wang et al., 2018) and includes one practice and one maximal effort trial per hand, with the order counter-balanced across participants. Handgrip endurance is measured by asking the participant to squeeze with 50% of their maximal effort using a target displayed through Acqknowledge for as long as they can tolerate. In order to index the physiological response to this mild stressor, beat-to-beat blood pressure is measured during the handgrip endurance test using an FDA approved Continuous Noninvasive Arterial Pressure system from CNSystems (Graz, Austria).

#### 1.1.4.2. Emotional Go/No-Go

Response inhibition has been assessed using a variety of different Go/No-Go tasks (Schulz et al., 2007; Wright et al., 2014). Response inhibition during emotional distraction is assessed using an emotional Go/No-Go task where participants are continuously presented with stimuli requiring either a button press (go), or no button press (no-go). 75 percent of trials are go trials, creating a tendency to go which must be inhibited for successful no-go trials. Positive, negative, and neutral images are presented behind the task-relevant stimuli in a manner adapted from (Cohen-Gilbert and Thomas, 2013) to measure the effect of emotional distraction.

#### 1.1.4.3. Computational Go/No-Go

We use a simplified version of the computational go/no-go task adapted from (Guitart-Masip et al., 2014) to measure learning under uncertainty. This task contains four cues, each indicating a different condition (go to win, go to avoid losing, no-go to win, no-go to avoid losing). Participants are instructed that they should figure out the best response for each stimulus, and that the best response may either be to go or no-go. After each response, participants are shown the outcome of the trial which can either be a win, loss, or no win/loss. Following a correct response, a favorable outcome occurs with 80 percent probability, and similarly, incorrect responses lead to unfavorable outcomes 80 percent of the time.

#### 1.1.4.4. Heartbeat Detection

The heartbeat detection task contains four trials where participants are asked to press a key in sync with either their heartbeat or a tone with varying instructions (Smith et al., 2020). In the first trial, participants are instructed to guess even when they cannot feel a heartbeat. The second trial consists of tones presented at approximately 80 tones per minute, and participants press a key in synchrony with the tones. On the third trial the instructions are to press along with their heartbeat, but only to press when certain about feeling a beat. The fourth trial is similar, except with the added instruction to complete the trial during an inspiratory breath hold. The breath-hold trial measures how much heartbeat awareness improves during interoceptive disturbance.

#### 1.1.4.5. Fear conditioning recall and recognition

In the recall phase of the fear conditioning task, participants are presented with the same stimuli (CS+ and CS-) as used during the scanning session, however, no US is presented. The recall phase contains 25 presentations of each stimulus as well as valence, arousal, and anxiety ratings. Recall of previously conditioned stimuli may be abnormal among people with anxiety disorders (Marin et al., 2017).

### 1.1.5. Biospecimens

Collection of biospecimens (blood, microbiome) allow us to assess molecular, genetic, and microbial levels of analysis. After initial processing, specimen storage is tracked using the Freezerworks inventory management system.

#### 1.1.5.1. Blood

Approximately 90mL of blood is collected via venipuncture using BD Vacutainer collection tubes for investigation of neuroendocrine, metabolic, inflammatory, cardiovascular, and genetic relationships with psychiatric illness. Samples are transferred to the University of Oklahoma Integrative Immunology Center (IIC) Laboratories, where initial processing, storage, and some assays take place. Plasma (from BD Vacutainer spray-coated K2EDTA tubes), serum (from BD Vacutainer silica spray-coated serum tubes), RNA (from PAXgene blood RNA tubes), and peripheral blood mononuclear cells (from BD Vacutainer Cell Preparation Tubes) are extracted and stored on-site, either at -80 °C or (for PBMCs) in liquid nitrogen dewars with monitors and alarms. Biospecimens are stored in multiple aliquots for ease of transfer to other laboratories and to ensure uninterrupted temperature conditions (avoidance of multiple freeze-thaw cycles). Once enough samples have been collected, they will either be further processed and analyzed in-house or sent to a contract laboratory for analyte quantification or genetic analysis.

#### 1.1.5.2. Microbiome

Participants provide microbial samples from the forehead, mouth, and stool using an all-in-one collection kit consisting of tubes labeled for body site, instructions, and cotton swabs. These samples are initially processed for DNA extraction using Qiagen DNeasy PowerSoil kits at IIC and stored, then later shipped to the University of California San Diego for final processing and analysis, e.g. 16s sequencing.

### 1.1.6. Actigraphy

Daily activity including step count, floors climbed, calories burned, heart rate and sleep are measured using FitBit Ionic devices over a 5-day period. Devices are issued to each participant with instructions not to remove the device until returning it. The FitBit Ionic and 5-day duration were chosen so that each assessment can be done without needing to recharge the battery or upload data until after the device is returned. After participants return the device, actigraphy data are imported into Fitabase for storage until later export.

Ader, D.N. (2007). "Developing the patient-reported outcomes measurement information system (PROMIS)". LWW).

Ball, T.M., Knapp, S.E., Paulus, M.P., and Stein, M.B. (2017). Brain activation during fear extinction predicts exposure success. *Depress Anxiety* 34(3)**,** 257-266. doi: 10.1002/da.22583.

Bartlett, M.S., Movellan, J.R., and Sejnowski, T.J. (2002). Face recognition by independent component analysis. *IEEE Trans Neural Netw* 13(6)**,** 1450-1464. doi: 10.1109/TNN.2002.804287.

Celis-Morales, C.A., Welsh, P., Lyall, D.M., Steell, L., Petermann, F., Anderson, J., et al. (2018). Associations of grip strength with cardiovascular, respiratory, and cancer outcomes and all cause mortality: prospective cohort study of half a million UK Biobank participants. *BMJ* 361**,** k1651. doi: 10.1136/bmj.k1651.

Cohen-Gilbert, J.E., and Thomas, K.M. (2013). Inhibitory control during emotional distraction across adolescence and early adulthood. *Child Dev* 84(6)**,** 1954-1966. doi: 10.1111/cdev.12085.

Donato, G., Bartlett, M.S., Hager, J.C., Ekman, P., and Sejnowski, T.J. (1999). Classifying Facial Actions. *IEEE Trans Pattern Anal Mach Intell* 21(10)**,** 974. doi: 10.1109/34.799905.

Ekman, P., Levenson, R.W., and Friesen, W.V. (1983). Autonomic nervous system activity distinguishes among emotions. *Science* 221(4616)**,** 1208-1210. doi: 10.1126/science.6612338.

Gerodimos, V., Karatrantou, K., Psychou, D., Vasilopoulou, T., and Zafeiridis, A. (2017). Static and Dynamic Handgrip Strength Endurance: Test-Retest Reproducibility. *J Hand Surg Am* 42(3)**,** e175-e184. doi: 10.1016/j.jhsa.2016.12.014.

Gershon, R.C., Wagster, M.V., Hendrie, H.C., Fox, N.A., Cook, K.F., and Nowinski, C.J. (2013). NIH toolbox for assessment of neurological and behavioral function. *Neurology* 80(11 Suppl 3)**,** S2-6. doi: 10.1212/WNL.0b013e3182872e5f.

Glover, G.H., Li, T.Q., and Ress, D. (2000). Image-based method for retrospective correction of physiological motion effects in fMRI: RETROICOR. *Magn Reson Med* 44(1)**,** 162-167. doi: 10.1002/1522-2594(200007)44:1<162::aid-mrm23>3.0.co;2-e.

Greenwald, M.K., Cook, E.W., and Lang, P.J. (1989). Affective judgment and psychophysiological response: dimensional covariation in the evaluation of pictorial stimuli. *Journal of psychophysiology*.

Guitart-Masip, M., Economides, M., Huys, Q.J., Frank, M.J., Chowdhury, R., Duzel, E., et al. (2014). Differential, but not opponent, effects of L -DOPA and citalopram on action learning with reward and punishment. *Psychopharmacology (Berl)* 231(5)**,** 955-966. doi: 10.1007/s00213-013-3313-4.

Hamilton, C.M., Strader, L.C., Pratt, J.G., Maiese, D., Hendershot, T., Kwok, R.K., et al. (2011). The PhenX Toolkit: get the most from your measures. *Am J Epidemiol* 174(3)**,** 253-260. doi: 10.1093/aje/kwr193.

Harris, P.A., Taylor, R., Minor, B.L., Elliott, V., Fernandez, M., O'Neal, L., et al. (2019). The REDCap consortium: Building an international community of software platform partners. *J Biomed Inform* 95**,** 103208. doi: 10.1016/j.jbi.2019.103208.

Harris, P.A., Taylor, R., Thielke, R., Payne, J., Gonzalez, N., and Conde, J.G. (2009). Research electronic data capture (REDCap)--a metadata-driven methodology and workflow process for providing translational research informatics support. *J Biomed Inform* 42(2)**,** 377-381. doi: 10.1016/j.jbi.2008.08.010.

Homma, I., and Masaoka, Y. (2008). Breathing rhythms and emotions. *Exp Physiol* 93(9)**,** 1011-1021. doi: 10.1113/expphysiol.2008.042424.

Knutson, B., Fong, G.W., Adams, C.M., Varner, J.L., and Hommer, D. (2001). Dissociation of reward anticipation and outcome with event-related fMRI. *Neuroreport* 12(17)**,** 3683-3687. doi: 10.1097/00001756-200112040-00016.

Licht, C.M., De Geus, E.J., Van Dyck, R., and Penninx, B.W. (2009). Association between anxiety disorders and heart rate variability in The Netherlands Study of Depression and Anxiety (NESDA). *Psychosomatic medicine* 71(5)**,** 508-518.

Lien, J.J., Kanade, T., Cohn, J.F., and Li, C.-C. (Year). "Automated facial expression recognition based on FACS action units", in: *Proceedings Third IEEE International Conference on Automatic Face and Gesture Recognition*: IEEE), 390-395.

Marin, M.F., Zsido, R.G., Song, H., Lasko, N.B., Killgore, W.D.S., Rauch, S.L., et al. (2017). Skin Conductance Responses and Neural Activations During Fear Conditioning and Extinction Recall Across Anxiety Disorders. *JAMA Psychiatry* 74(6)**,** 622-631. doi: 10.1001/jamapsychiatry.2017.0329.

Miu, A.C., Heilman, R.M., and Miclea, M. (2009). Reduced heart rate variability and vagal tone in anxiety: trait versus state, and the effects of autogenic training. *Auton Neurosci* 145(1-2)**,** 99-103. doi: 10.1016/j.autneu.2008.11.010.

Najstrom, M., and Jansson, B. (2006). Unconscious responses to threatening pictures: interactive effect of trait anxiety and social desirability on skin conductance responses. *Cogn Behav Ther* 35(1)**,** 11-18. doi: 10.1080/16506070510011566.

Sanislow, C.A., Pine, D.S., Quinn, K.J., Kozak, M.J., Garvey, M.A., Heinssen, R.K., et al. (2010). Developing constructs for psychopathology research: research domain criteria. *Journal of abnormal psychology* 119(4)**,** 631.

Schulz, K.P., Fan, J., Magidina, O., Marks, D.J., Hahn, B., and Halperin, J.M. (2007). Does the emotional go/no-go task really measure behavioral inhibition? Convergence with measures on a non-emotional analog. *Archives of Clinical Neuropsychology* 22(2)**,** 151-160.

Simmons, W.K., Avery, J.A., Barcalow, J.C., Bodurka, J., Drevets, W.C., and Bellgowan, P. (2013). Keeping the body in mind: insula functional organization and functional connectivity integrate interoceptive, exteroceptive, and emotional awareness. *Hum Brain Mapp* 34(11)**,** 2944-2958. doi: 10.1002/hbm.22113.

Smith, R., Kuplicki, R., Feinstein, J., Forthman, K.L., Stewart, J.L., Paulus, M.P., et al. (2020). An active inference model reveals a failure to adapt interoceptive precision estimates across depression, anxiety, eating, and substance use disorders. *medRxiv*.

Susskind, J.M., Littlewort, G., Bartlett, M.S., Movellan, J., and Anderson, A.K. (2007). Human and computer recognition of facial expressions of emotion. *Neuropsychologia* 45(1)**,** 152-162. doi: 10.1016/j.neuropsychologia.2006.05.001.

Wang, Y.C., Bohannon, R.W., Li, X., Sindhu, B., and Kapellusch, J. (2018). Hand-Grip Strength: Normative Reference Values and Equations for Individuals 18 to 85 Years of Age Residing in the United States. *J Orthop Sports Phys Ther* 48(9)**,** 685-693. doi: 10.2519/jospt.2018.7851.

Watkins, L.L., Grossman, P., Krishnan, R., and Sherwood, A. (1998). Anxiety and vagal control of heart rate. *Psychosom Med* 60(4)**,** 498-502. doi: 10.1097/00006842-199807000-00018.

Wright, L., Lipszyc, J., Dupuis, A., Thayapararajah, S.W., and Schachar, R. (2014). Response inhibition and psychopathology: a meta-analysis of go/no-go task performance. *J Abnorm Psychol* 123(2)**,** 429-439. doi: 10.1037/a0036295.

Wu, T., Butko, N.J., Ruvolo, P., Whitehill, J., Bartlett, M.S., and Movellan, J.R. (2012). Multilayer architectures for facial action unit recognition. *IEEE Transactions on Systems, Man, and Cybernetics, Part B (Cybernetics)* 42(4)**,** 1027-1038.
